# Recommendations for developing asynchronous online consultations for chlamydia treatment for underserved populations: A Behaviour Change Wheel analysis

**DOI:** 10.1101/2025.07.08.25330192

**Authors:** Claudia S. Estcourt, Julie McLeod, Paul Flowers, Jennifer MacDonald, Fiona Mapp, John Saunders, Melvina Woode Owusu, Amelia McInnes-Dean, Nuria Gallego Marquez, Ann Blandford, Pam Sonnenberg, Jo Gibbs

**Affiliations:** School of Health and Life Sciences, Glasgow Caledonian University, Glasgow, Scotland, United Kingdom; Psychological Science and Health, University of Strathclyde, Glasgow, Scotland, United Kingdom; Institute for Global Health, University College London, London, England, United Kingdom; UK Health Security Agency (UKHSA), London, England, United Kingdom; UCL Interaction Centre (UCLIC), University College London, London, England, United Kingdom

**Author notes:** **Corresponding author:** Claudia S. Estcourt.

## Abstract

**Introduction:** People from underserved groups experience disproportionately poor sexual health and challenges accessing care. Asynchronous online consultations (a user completes a health questionnaire online, which is reviewed by a clinician) are being used within sexual healthcare to prescribe chlamydia treatment. Users require sufficient health and digital literacy to access online services and use them safely.

**Methods:** We used the PROGRESS-Plus framework to guide purposive recruitment of 35 participants from diverse underserved groups, from community settings and sexual health services in contrasting areas of the UK (15/10/21-18/03/22). We conducted qualitative semi-structured interviews and thematic analyses to derive key barriers and facilitators to using asynchronous online consultations. We used the Behaviour Change Wheel to specify recommendations to address them.

**Results:** Over half of participants were from the most deprived areas and 40% were from minoritised ethnic groups. Key barriers included: lack of familiarity with online healthcare; perceived need to see a healthcare professional in person; privacy concerns; concerns about difficulty interpreting the questions; discomfort answering personal questions online. Key facilitators included: familiarity with online consultations; perceived low sexually transmitted infection (STI) risk; perceived increase in convenience, control, and privacy; simple wording and design; and support whilst completing them. Recommendations included: increasing awareness and familiarity by promoting them offline and online, and providing demonstrations and instructions on how to use them; encouraging people to choose them by highlighting available support, equivalence to in-person consultations, and privacy and convenience; and reducing attrition by using simple wording and design, providing additional explanations, and offering audio and visual alternatives to text.

**Conclusions:** Incorporating these evidence-based, theoretically informed recommendations could widen access to underserved groups, and increase the usability and safety of asynchronous online consultations for chlamydia treatment. Recommendations are likely to benefit all users and could be of use across health more broadly.

**Main Messages:**

1. Some sexual health services provide chlamydia treatment through asynchronous online consultations, where patients complete an online health questionnaire which is reviewed by a clinician before issuing a prescription, without the need for direct interaction with a healthcare professional.
2. Safe and effective use of asynchronous online consultations requires adequate digital and health literacy but there is limited evidence on how to design these consultations inclusively, posing a risk of excluding underserved groups and exacerbating health inequalities.
3. Among people from underserved groups, we found key barriers to using asynchronous online consultations included: a lack of familiarity with online healthcare; perceived need to see a healthcare professional in person; privacy concerns; concern about difficulty interpreting the questions; discomfort answering personal questions online.
4. Key facilitators included: familiarity with online consultations; perceived low STI risk; perceived increase in convenience, control, and privacy; simplicity in consultation wording and design; and support whilst completing them.
5. Recommendations include: increasing awareness and familiarity by promoting asynchronous online consultations offline (e.g. in GP practices) and online (e.g., social media), and providing demonstrations/instructions on how to use them; encouraging uptake by highlighting options for support, equivalence to in-person consultations, and privacy and convenience; and reducing attrition by using simple wording and design, providing additional explanations, and offering audio/visual alternatives to text.

## Introduction

Health inequalities persist globally, disproportionately affecting underserved populations who face significant barriers to timely and appropriate healthcare. These disparities are particularly evident in sexual health, where access to prevention, testing, and treatment services remains uneven.^1 2^ Underserved groups/populations, defined as those with inadequate healthcare access and underrepresentation in research,^3^ include sex workers, people experiencing homelessness, those involved with the criminal justice system, individuals with substance dependence, trans and gender-diverse people, vulnerable migrants, victims of modern slavery, people with learning disabilities, severe mental illness, physical disabilities, and those with low digital and health literacy.^4^ These populations may experience disproportionately poor sexual health outcomes, including higher rates of STIs and delayed access to care and treatment.^4^

In some countries, sexual health services increasingly include a range of online services such as STI and blood-borne virus testing via online postal self-sampling,^1 5^ or by requesting testing online, followed by sample collection at a laboratory.^6^ More recently, online asynchronous consultations have been introduced to manage infections, such as chlamydia.^7^ This involves a patient diagnosed with chlamydia completing an online health questionnaire, which is then reviewed by a clinician who prescribes antibiotic treatment (if safe to do so) based on the information provided, without needing direct interaction with the patient.

While asynchronous consultations could reduce barriers to in-person healthcare, they could also exacerbate existing health inequalities or create new ones. Many underserved individuals, including those with low digital and health literacy may face structural barriers to accessing online services such as lack of connectivity and/or software access.^8^ Others could (also) struggle to navigate online health questionnaires effectively, limiting access to necessary care and/or their safe use. Safe prescribing requires adequate knowledge of the patient^9^ and while many regulatory bodies provide guidance on remote prescribing,^9^ there is little empirical evidence to inform the content and design of the asynchronous online consultations on which the prescribing decision is based. Further, being underserved or experiencing multiple disadvantage can have a cumulative impact, where intersectional identities contribute to compounded disparities in access and health outcomes. Recent literature underscores the interplay between socio-demographic factors and digital healthcare access, highlighting how online consultations may unintentionally exclude those most in need.^10-12^ The usability and accessibility of online questionnaires within asynchronous online consultations are modifiable but despite the growing adoption of asynchronous consultations, there is limited evidence on how to design them inclusively for underserved populations.^13^

To address this gap, our study aimed to: i) identify barriers and facilitators to accessing chlamydia treatment via asynchronous online consultations among underserved populations and ii) develop evidence-based recommendations to overcome these barriers. The work will inform the optimisation of an inclusive eSexual Health clinic^14^ for a future trial.^15^

## Methods

### Participants

We developed a target sampling frame (Appendix A) using the PROGRESS-Plus framework^16^ to purposively recruit people from a range of underserved populations, stratified by inequalities in health opportunities and outcomes (Table 1). In line with Braun and Clarke (2006),^17^ we did not seek to meet data saturation. Instead, prior to recruitment, a sample of 35 was identified as appropriate to meet the sample targets and sufficient Information Power.^18^ After 35 interviews, we reviewed the data and were satisfied that Information Power had been attained.

**Table 1:** Participant self-reported socio-economic demographic characteristics.

| **Variables** | **n** | **% \*** |
| --- | --- | --- |
| ***Age*** |  |  |
| 16-24 | 8 | 22.9 |
| 25-34 | 10 | 28.6 |
| 35-44 | 13 | 37.1 |
| 45-54 | 1 | 2.9 |
| 55-64 | 1 | 2.9 |
| 65+ | 1 | 2.9 |
| ***Ethnicity\*\**** |  |  |
| Asian, Asian British or Asian Welsh: Chinese | 1 | 2.9 |
| Asian, Asian British or Asian Welsh: Pakistani | 4 | 11.4 |
| Black, Black British, Black Welsh, Caribbean or African: African | 3 | 8.6 |
| Mixed or Multiple ethnic groups: Other Mixed or Multiple ethnic groups | 1 | 2.9 |
| White: Irish | 1 | 2.9 |
| White: English, Welsh, Scottish, Northern Irish, or British | 19 | 54.3 |
| Other self-identified groups (e.g., Hungarian, Italian, Jewish) | 5 | 14.3 |
| ***Sexual orientation*** |  |  |
| Bisexual (M) | 2 | 5.7 |
| Bisexual (F) | 5 | 14.3 |
| Bi/pansexual/queer (gender diverse) | 3 | 8.6 |
| Heterosexual/straight (M) | 6 | 17.1 |
| Heterosexual/straight (F) | 10 | 28.6 |
| Gay (M) | 7 | 20.0 |
| No response | 1 | 2.9 |
| ***Gender*** |  |  |
| Cisgender woman | 16 | 45.7 |
| Cisgender man | 16 | 45.7 |
| Gender diverse (non-binary, trans masc) | 2 | 5.7 |
| ***Education (n=33)*** |  |  |
| Secondary/High school | 6 | 17.1 |
| College (introductory/foundational vocational award to diploma) | 12 | 34.3 |
| University (Undergraduate student) | 5 | 14.3 |
| University (Undergraduate degree achieved) | 5 | 14.3 |
| University (Postgraduate degree achieved) | 5 | 14.3 |
| No response | 1 | 2.9 |
| **Occupation** |  |  |
| Unemployed | 11 | 31.4 |
| Student (full-time/part-time) | 9 | 25.7 |
| Employed (full-time/part-time) | 13 | 37.1 |
| Retired | 1 | 2.9 |
| **Area of deprivation\*\*\*** |  |  |
| IMD Quintile 1 (most deprived) | 9 | 25.7 |
| IMD Quintile 2 | 8 | 22.9 |
| IMD Quintile 3 | 6 | 17.1 |
| IMD Quintile 4 | 2 | 5.7 |
| IMD Quintile 5 (least deprived) | 6 | 17.1 |
| No postcode (i.e., homeless) | 1 | 2.9 |
| No response | 3 | 8.6 |
| **Difficulty making ends meet\*\*\*\*** |  |  |
| Yes | 9 | 25.7 |
| Sometimes | 3 | 8.6 |
| Not anymore but have in the past | 2 | 5.7 |
| No | 20 | 57.1 |
| **Belong to any particular religion or faith** |  |  |
| No (never, not anymore) | 20 | 57.1 |
| Maybe (e.g., spiritual, agnostic) | 2 | 5.7 |
| Yes | 12 | 34.3 |
| Christianity | 7 | 20.0 |
| Islam | 4 | 11.4 |
| Judaism | 1 | 2.9 |
| **Disability** |  |  |
| Learning disability (n=34) | 10 | 28.6 |
| Mental or physical disability (n=33) | 17 | 48.6 |
| *Reduces ability to carry out day-to-day activities* | 12 | 34.3 |
| *Sometimes reduces ability to carry out day-to-day activities* | 1 | 2.9 |
| *Does not reduce ability to carry out day-to-day activities* | 3 | 8.6 |
| **First language** |  |  |
| English | 26 | 74.3 |
| Non-English (Italian, Indonesian, French, Hungarian, Yoruba, Urdu, Gaelic) | 8 | 22.9 |
| **Country born** |  |  |
| UK (England, Scotland) | 26 | 74.3 |
| *England* | 13 | 37.1 |
| *Scotland* | 13 | 37.1 |
| Non-UK (Italy, Indonesia, Switzerland, Hungary, Nigeria) | 8 | 22.9 |

**Legend:** \*Participant demographics for one participant were not obtained, table includes demographics for n=34, except where participants did not wish to answer the question. Percentages are calculated for N=35; those not summing to 100 are due to rounding. \*\*Reported using the Official National Statistics classifications <sup>24</sup>. \*\*\*Index of Multiple Deprivation.<sup>22</sup> \*\*\*\*“Making ends meet” = ability to pay for essentials needed to live.

### Recruitment

To reach a diverse range of participants, we collaborated with four NHS Trusts in England and one NHS Board in Scotland - large, state funded healthcare organisations, along with three non-governmental organisations (NGOs). These NGOs included one supporting people with learning disabilities, one serving lesbian, gay, bisexual, transgender, queer and intersex+ (LGBTQI+) people, and one focused on Muslim community members. Additionally, we partnered with one college (for national diplomas and qualifications) in a deprived area of Scotland, serving people with low educational attainment.

Representatives from each NHS Trust/Board, NGO and college identified potential participants who appeared to meet the inclusion criteria and with permission, referred them to the researcher (JMcL). JMcL conducted a 15-minute phone survey (Appendix A) to check eligibility, collect socio-economic demographic information, evaluate technology access and digital confidence, address queries and schedule the interviews. Participants were then provided with a consent form, participant information sheet, and interview visual aids (Figure 1) to read, either by email, WhatsApp, or hard copy.

**Figure 1:**
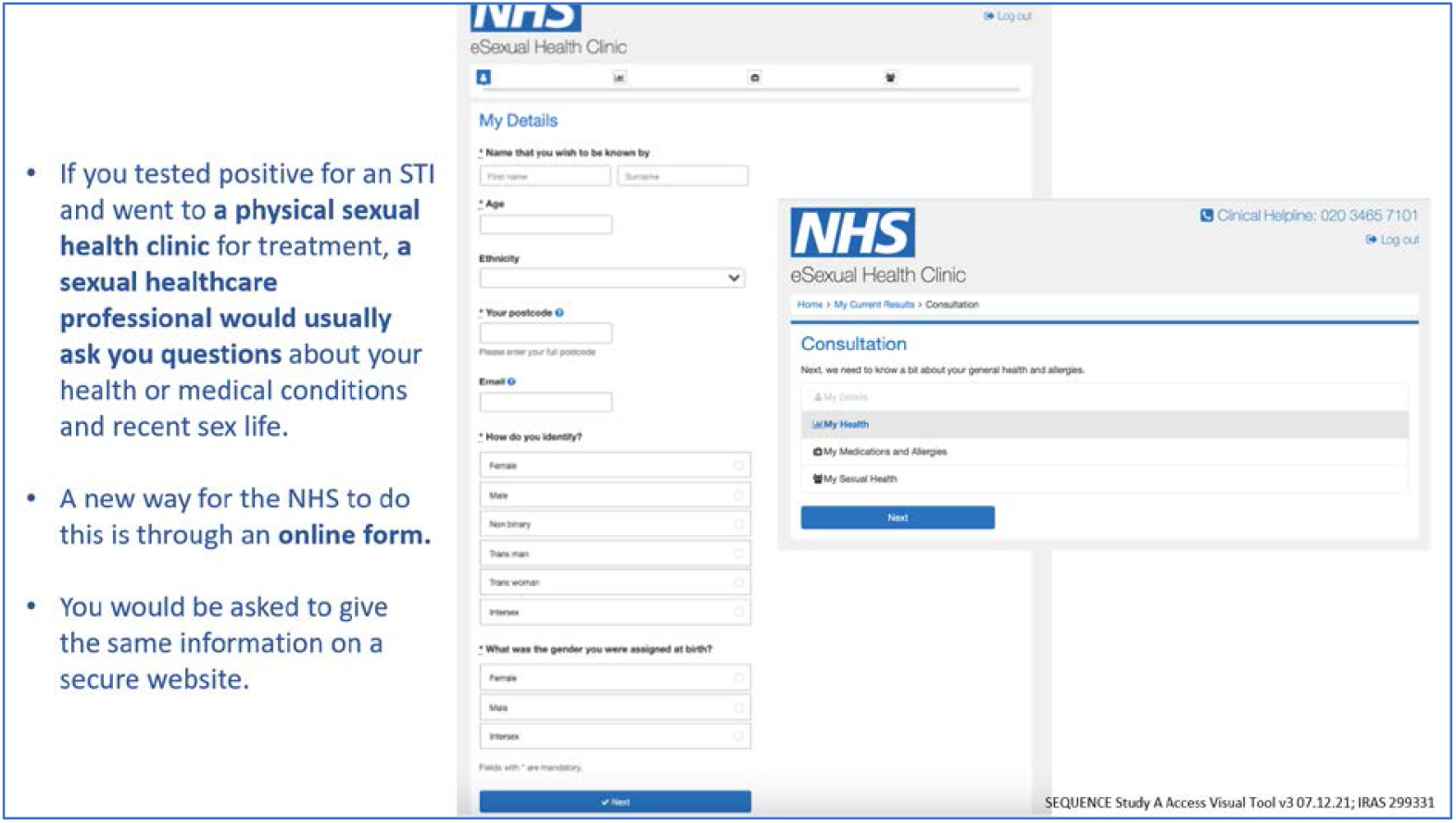
Examples of visual aids used as prompts to collect focussed data regarding asynchronous online consultation. **Legend:** Slides represented typical screenshots from our eSexual Health Clinic prior to optimisation

Inclusion criteria were: aged 16 years and older; resident in the UK; access to the internet and a telephone; able to read and speak English well enough to participate; sexually active; and never used, or struggled to use, current postal STI self-sampling services.

Throughout recruitment, we monitored the sample closely by checking the demographics of potential participants against the existing sample and target sampling frame. As sample targets were met, we liaised with NHS, NGO and college contacts to focus recruitment of participants with characteristics not already included.

### Procedure

JMcL conducted interviews 15/10/21-18/03/22, either remotely via phone call (n=23) or video call using Microsoft Teams/Zoom (n=7), or face-to-face (n=5). Consent was obtained at the beginning of each interview, verbally on an encrypted audio recorder for remote interviews, and written for face-to-face interviews. Interviews were semi-structured (Appendix A: interview topic guide) ranging from 38-82 minutes duration (*median*=60), focussing on participant-led data collection by prompting participants to elaborate on their responses. Throughout, participants were referred to visual aids (Figure 1) and regular sense checks were conducted to ensure the participants had a clear understanding of the asynchronous online consultation. At the end of the interviews, all participants were offered a shopping voucher worth £30 and provided with sexual health support resources (Appendix A).

All interviews were audio recorded using encrypted digital devices; recordings were transcribed verbatim for analysis. Transcripts were fully anonymised for reporting, presentation, and publication purposes.

### Analysis

JMcL conducted a two staged analysis under the supervision of PF. In Stage one, using NVivo (version 12), we conducted inductive thematic analysis^17^ to derive participant-led barrier and facilitator themes. We initially described data with brief summary barrier and facilitator statements which we then grouped with similar summary statements to identify subsequent themes and subthemes. We resolved any disagreements on the naming and grouping of statements, themes, and subthemes through discussion.

In Stage two, we used the Behaviour Change Wheel^19^ to develop recommendations to overcome the barriers and enhance the facilitators. First, we coded barrier and facilitator themes to the Theoretical Domains Framework.^20^ We then matched these domains with corresponding intervention functions and further operationalised them with specific behaviour change techniques (BCTs) using the BCT Taxonomy version 1^21^ to provide evidence-based and theoretically informed recommendations. Clinicians CSE, JG, JS, PS reviewed the recommendations for clinical relevance.

### Patient and public involvement

For material development, we consulted 12 public and patient involvement and engagement (PPIE) representatives of diverse ages, genders, ethnicities, sexual orientations, religions, and experiences of disability, learning difficulties, and digital STI healthcare. Five representatives offered intersectional perspectives and advice on our questionnaire-based assessment of participant demographics and internet use, and seven advised on the interview topic guide and visual aids to be used within data collection.

### Ethical considerations

Ethical approval for this study was granted by the East of England - Cambridge South Research Ethics Committee (REC) (reference 21/EE/0148) and Glasgow Caledonian University REC (reference HLS/NCH/20/045).

## Results

### Participants

The 35 participants (Table 1) (age range 18-70 years, *mean*=34) came from various underserved populations; for example, 18 (51%) lived in the most deprived areas of the UK (defined by the Index of Multiple Deprivation)^22^ and one (3%) was experiencing homelessness; 18 (51%) had no higher (post-college) education; 15 (40%) were of a ethnic minority group,^23^ eight (23%) reported English was not their first language; 17 (49%) had a mental health or physical illness or condition lasting 12 months or more; and 10 (29%) had a learning difficulty.

Participants used the internet extensively (mainly for social media) and self-rated their online skills as high. However, amongst the examples of internet activities reported, very few included using the internet for more sophisticated tasks, such as online banking, and few had experience of online healthcare (Appendix A).

Six main barrier and facilitator themes were identified from the inductive analysis (Stage 1): awareness and familiarity; perceived needs; convenience and resources; privacy and disclosures, answering questions correctly; and answering personal questions online. These are shown in Table 2 along with indicative quotations from participants. The BCW analysis (Stage 2) built on the barrier and facilitator themes to provide systematic recommendations to enhance the use of online consultations are shown in Table 3. Please see Appendix B for the full BCW analysis.

**Table 2:** Overview of barrier and facilitator themes to using asynchronous online consultations for chlamydia treatment.

| Barrier | Facilitator | Indicative quotes |
| --- | --- | --- |
| People find it <u>hard</u> to do an asynchronous online consultation... | People find it <u>easy</u> to do an asynchronous online consultation... |  |
| **Theme 1: Awareness and familiarity** |  |  |
| **...if they lack familiarity with online healthcare**<br>Participants reported a lack of awareness and experience with online sexual healthcare and a concern about doing it wrong and needing to practise due to their inexperience. | **...if they have experience with online forms or can familiarise themselves with the online consultation**<br>Participants reported the benefit of having previous experience completing online forms or having the option to become familiar with the online consultation before attempting to complete it (e.g., see the questions, see a completed example, practise first). | *"I don't think...just to fill that in, I don't think it would be much of a challenge apart from if it's simple to do, doing online things. If it's not...I'd have to get one through to try it to see if it was challenging or not."* |
| **Theme 2: Perceived needs** |  |  |
| **...if they perceive the need to see a healthcare professional in person**<br>Some felt in-person interactions were necessary so that they could be examined particularly if they perceived themselves to be at greater likelihood of having an STI for example if they had symptoms. Others wanted an in-person interaction to have questions answered. | **...if their perceived level of risk of an STI is low**<br>Participants were happy to complete the online consultation instead of seeing a healthcare professional (HCP) in-person if they perceived themselves to be "at low risk", i.e., they do not have any symptoms. | *"Well, I guess, at that point, I wouldn't use the online. So, if...yeah, after being tested positive for any sort of STI, I would want to actually have a real person in front of me [...] because of how impersonal it feels, and now that it's [...] questions about something that has been confirmed professionally, I think I would want to actually, yeah, talk to someone real."* |
| **Theme 3: Convenience and resources** |  |  |
| **No theme identified** | **...if they perceive it to increase convenience and control**<br>Participants perceived that an online consultation is more convenient than in-person and is potentially faster and more time efficient than in person. | *"maybe you take...you need to take a bus, waiting for a bus, and staying on the bus for maybe 20 minutes/ 30 minutes to get to the hospital or to get to the centre where you would take some time, that would be... it maybe a lot of stress on it. But doing... filling your form online and following it up is... something like this easier for me. Yeah."* |

| Theme 4: Privacy and disclosures |  |  |
| --- | --- | --- |
| **...if they are concerned about others reading their responses**<br>Participants expressed concerns about others, such as parents or partners, finding out they had an STI and required treatment | **...if it is perceived it to be private**<br>Perception that the online consultation was private and they would remain confidential was important. The online consultation was also perceived to offer protection from negative experiences in clinic/with healthcare professionals, such as judgement or stigma, or concern about safety and comfort (e.g., Muslim females with male healthcare professionals), and for some participants, an asynchronous online consultation could make them feel comfortable in reporting more behaviours which put them at higher risk of STIs. | *"I think that online consultation, that form...if that would have been there when I was on the chem sex, so with the depression and that, I probably wouldn't have HIV, because I'd have openly admitted that I injected drugs into my system, and sometimes I'm so impaired I don't know, and I would have openly admitted it. But when you're face-to-face with someone, you won't admit it. Anyone that I know, [but I'm not saying], has said they never admitted it at the sexual health centre, they'd never admit it"* |
| Theme 5: Answering questions correctly |  |  |
| **...if they have difficulty understanding or interpreting the questions**<br>Participants expressed concern about difficulty reading, misunderstanding or misinterpreting the questions, particularly those about their health (rather than sexual behaviour), for fear of getting incorrect treatment. | **...if the layout and questions of the consultation are simple and clear or they have support to complete it**<br>Participants expressed the need for the look and flow of the online consultation and the questions to be straightforward, simple and clear (i.e., be step-by-step), for extra information and pictures to help self-identify health issues and for the form to be in easy-read format with pictures. Some also felt that that having options for support from a healthcare professional would be necessary. | *"I think with the symptoms, it would be good to have information so you could be more specific with it, like specify what those things are, in case it's unclear, like 'bleeding between your periods', that could mean you're spotting a couple of days after your period or a couple of days before it or it could mean heavy bleeding between periods, that kind of thing. [...] So, just have the option to click on it to get more specific information on what that should include with you clicking on that option."* |
| Theme 6: Answering personal questions online |  |  |
| **...if they are uncomfortable answering personal questions**<br>Participants reported that they would feel discomfort answering personal questions, such as those about their gender, sexuality, or sexual health history, due to concerns about discrimination, judgement or loss of anonymity, gender questions being cis-normative, or | **...if the questions are perceived to be necessary and expected**<br>Participants reported the importance of knowing that the questions would be the same as would be asked in person, and necessary to get correct treatment. | *"I don't know whether, from a religious perspective, like, people might be afraid? [...] So, there is that religious setback I think a lot of people have coming from conservative households and coming from a conservative religion where they feel like they'd probably be letting God down by putting information out there. I know my sister's specific sect of Islam, they're not allowed to discuss sexual matters outside of their marriage and people aren't allowed to know. So, that could be a thing that kind of holds her back because filling in this form, she's probably* |
| discussing sexual health with anyone contradicting their faith. |  | *thinking, 'I'm letting other people know what's happening', as opposed to 'I'm looking for help'."* |

**Table 3:**
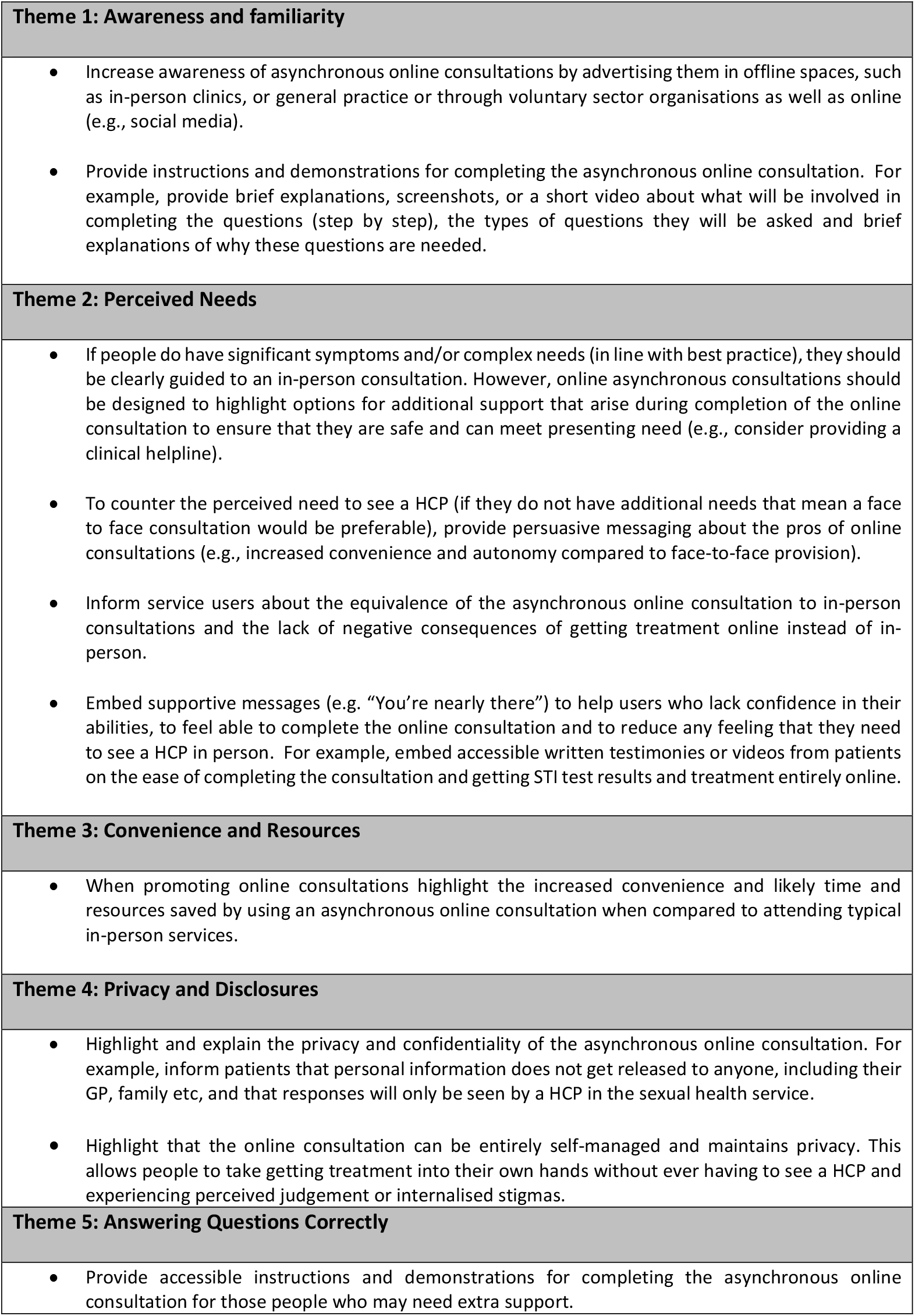
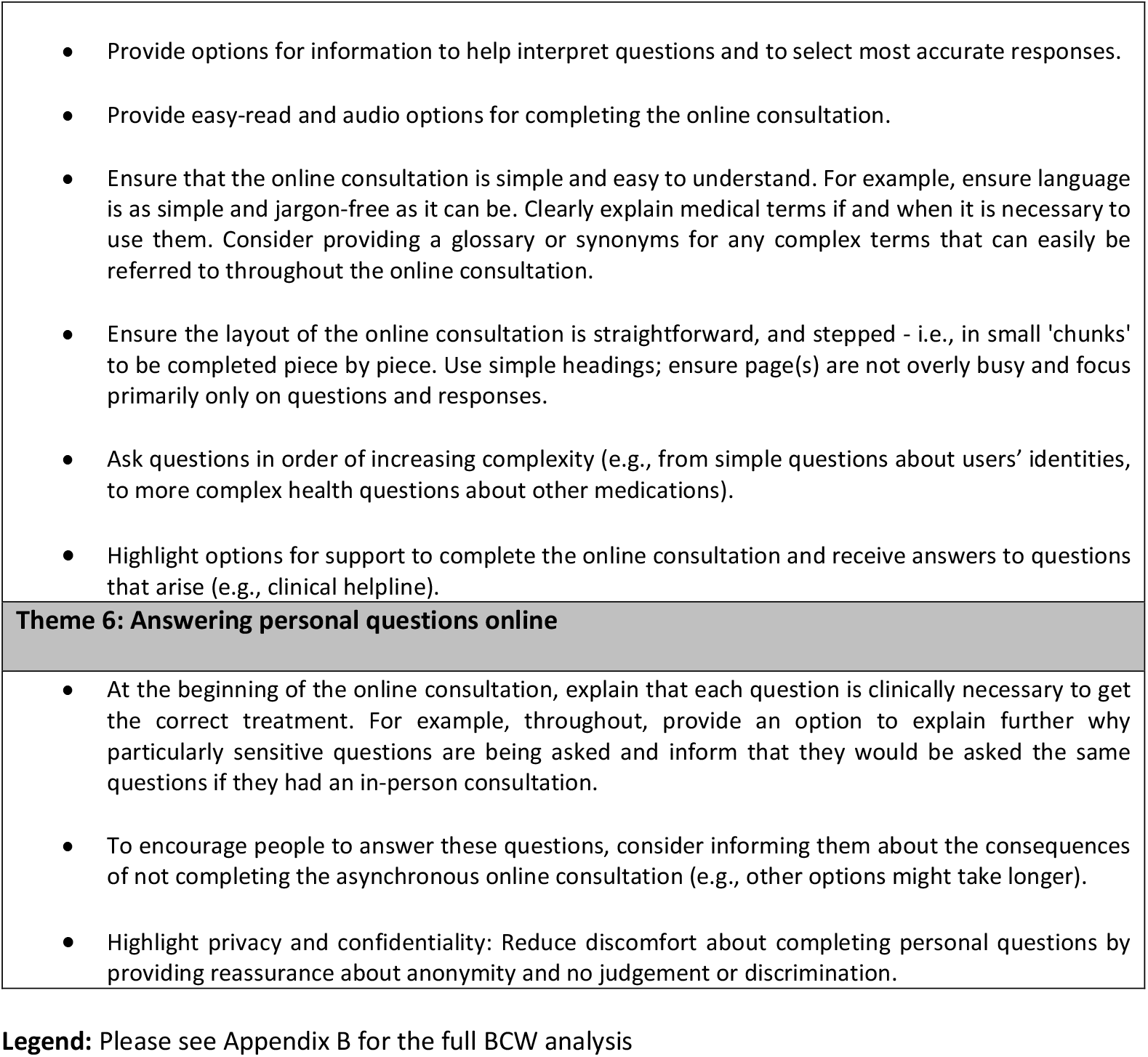
Recommendations to overcome the barriers and enhance the facilitators to using online asynchronous consultations, derived from the BCW analysis and grouped according to theme.

## DISCUSSION

To our knowledge, this is the first study to describe, evidence based and theoretically informed recommendations for developing online asynchronous consultations, which also focus on the needs of people from underserved groups. Key barriers to using asynchronous online consultations included: a lack of familiarity with online healthcare; perceived need to see a healthcare professional in person; privacy concerns; concerns about difficulty interpreting the questions; and discomfort answering personal questions online. Key facilitators included: familiarity with (any) online consultations; perceived low risk of having an STI; perceived increase in convenience, control, and privacy; simple wording and design; and support whilst completing them. Recommendations include; increasing awareness and familiarity by promoting them offline and online, and providing demonstrations/instructions on how to use them; encouraging people to choose them by highlighting available support, equivalence to in-person consultations, and privacy and convenience; and reducing attrition by using simple wording and design, providing additional explanations, and offering audio/visual alternatives to text.

We previously proposed a framework for developing the content and flow of medical questions in online asynchronous consultations^25^ but this did not explicitly address the needs of underserved groups. Although one study provides recommendations for implementation of asynchronous telemedicine platforms into primary care,^13^ few studies have focussed on the digital health needs of underserved groups and none have specifically explored issues relating to asynchronous online consultations leading to prescription of treatment, nor focussed specifically on sexual health care. Some countries have national accessibility guidelines^26^ and broad principles relating to digital exclusion but these lack the granularity needed. Our findings address this gap.

A systematic review of asynchronous telemedicine consultations in general practice concluded that they could increase access to timely care, although equity was poorly reported in the included studies.^10^ A systematic review of characteristics of “eConsultation” users in primary care reported that socio-demographically disadvantaged users used eConsultations less than other groups.^12^ Other studies report that HCPs perceive that online consultations could differentially disadvantage people who experience digital exclusion.^11 13 27^ However, it is unclear whether any of the systems used were developed with content/design tailored to those with lower digital and or health literacy, which could mitigate some access issues.

Similar to the findings of this study, Leighton et al’s analysis of the implementation of online asynchronous telemedicine platforms into primary care, found that patients (and staff) were concerned about privacy and confidentiality.^13^ Recommendations from a behaviour change wheel analysis of barriers and facilitators to seeking online sexual health information and support among underserved populations, derived from the same interviews as in this study, also found that online and offline promotion and endorsement by healthcare professionals and peers would reduce barriers to uptake.^28^

A study of users of HIV Pre-Exposure Prophylaxis (PrEP) in Scotland (participants were all men who have sex with men) suggested that some people would find it easier to provide accurate details about current medication when doing so online, as they would not feel time-pressured and could check medication details at home.^29^ Overall, these findings highlight the timeliness and importance of the current study for ensuring that inequalities, both digital and longstanding, are not widened by the shift to delivering healthcare online.

By working closely with NGOs and clinical services, and using targeted recruitment, we included the perspectives of people from groups who are seldom heard and often underserved, including people with learning disabilities. However, we missed target recruitment for gender diverse people, limiting insights for this group. We did not seek to sample or analyse data in relation to intersectionality but recognise that intersectional demographics and experiences can put people at even greater risk of poor sexual health and result in additional barriers to accessing care.^30^ Our methods extended beyond thematic analysis to the use of wider, robust behavioural science tools (the BCW) for systematic development of recommendations built both on the evidence we collected and on previous theory and cumulative knowledge (Appendix B).

Few participants had used online health care, so it was hard for some to conceptualise the eSexual Health Clinic. While we attempted to mitigate this by using screenshots (Fig 1) and careful explanations, some participants did not have a precise understanding of all the features of the eSexual Health Clinic which limited the specificity of their barriers and facilitators. Further, although digital and health literacy tools were available,^31^ we developed our own survey (Appendix A) to avoid inadvertently excluding people from the research due to difficulties completing the assessment tool. This may have complicated the recruitment of participants of lower digital and health literacy, as indicated by the number of participants who self-reported as having ‘high’ skills using the internet when their reported online activity suggested less complex usage.

Recommendations from this study could be used in conjunction with appropriate broader national guidance by service planners and providers to develop novel, and improve existing, asynchronous online consultations for maximum inclusivity. Many of the recommendations, such as those to increase awareness and help users complete the consultations could be straightforward to implement and beneficial for all users. Techniques from Human–Computer Interaction (HCI) should also be used in conjunction with our recommendations to ensure that the digital component of the intervention is efficient, effective and provides a positive user experience.^32^ Key stakeholders should thoroughly consider local factors in implementation to ensure recommendations are meaningful and appropriate in each setting, and to ensure asynchronous online consultations are integrated into existing systems and work flows.^13^

We have used the findings from this study to optimise our eSexual Health Clinic^14^ for maximal inclusivity and reach, ahead of a forthcoming randomised controlled trial.^15^ We will include an integral health inequalities evaluation as well as a distributional cost effectiveness analysis to evaluate the extent to which our design has influenced known differences in uptake and use of online sexual health systems, according to sociodemographic and other characteristics. Trials such as this, with a clear equity focus, are needed to fill the evidence gaps^10^ and ensure digital health meets its potential to narrow, rather than widen, health inequalities.

Uptake of these recommendations could increase access to, and successful completion of, asynchronous online consultations for all populations, including those who are underserved. This, in turn, could reduce inequalities in health outcomes. Although developed in the context of a clinical consultation for treatment of an STI, we believe that the recommendations are broadly generalisable across the population and for use with other health conditions.

By directly focussing on people from underserved groups, the findings from this study could improve existing online asynchronous consultation systes and inform the development of new ones for maximum inclusivity. The findings could be easily applied across online health care for different health conditions, particularly when designing with a focus on the needs of underserved groups. It is likely that all users, not just the underserved, would benefit from the simplicity, clarity, and usability features suggested by the participants. However, as remote service access is not always appropriate, nor accessible due to non-modifiable factors, they must be provided alongside accessible, non-digital, alternatives.

## Supporting information

GUIDED checklist

## Data Availability

Due to the sensitive nature of the questions asked in this study, participants were assured that transcripts would remain private and confidential and would not be shared beyond the use of anonymised illustrative quotes in publications about the research. In line with our ethical approvals form Glasgow Caledonian University (GCU) Nursing and Community Health Research Ethics Committee (HLS/NCH/20/045) and the East of England - Cambridge South Research Ethics Committee (21/EE/0148), participants were not asked and did not consent to sharing their full transcripts. The transcripts will be stored long-term, for a minimum of 10 years (from 2021) on a secure GCU network drive. Data access requests may be made to the corresponding author, Claudia Estcourt, the SEQUENCE digital team or the Glasgow Caledonian University Research Ethics Committee and the East of England - Cambridge South Research Ethics Committee.

## Acknowledgements

We would like to thank the participants who took part in the study. We are also grateful to the following sexual health services and organisations for invaluable assistance with the study: Castle Circus Health Centre (Torbay & South Devon NHS Foundation Trust), Devon Sexual Health – Exeter & Barnstaple (Northern Devon NHS Foundation Trust), Mortimer Market Centre, Archway, Barnet and Buryfields Clinics (Central and North West London NHS Foundation Trust), Sandyford Clinic (Greater Glasgow & Clyde), and Sexual Health Sheffield (Sheffield Hospitals NHS Foundation Trust) for advertising the study to support with recruitment. We are very grateful to our sponsor, Central and North West London NHS Foundation Trust, and the staff of the Noclor NHS Research team for their support and assistance. We would also like to thank the representatives of Hidayah LGBT, get2gether, and Scottish Wider Access Programme (SWAP) college courses for publicising the study to help with recruitment.

Finally, we would like to thank the members of our Community Advisory Panel whose valuable input supported the development of an inclusive participant recruitment strategy and accessible, culturally appropriate data collection materials (consent forms, interview materials, demographic questionnaires, topic guides, case scenarios and clinical pathway diagrams).

This study/project is funded by the NIHR Programme Grants for Applied Research, NIHR200856. The views expressed are those of the author(s) and not necessarily those of the NIHR or the Department of Health and Social Care.

*SEQUENCE Digital team: Claudia Estcourt (PI), Vanessa Apea, Ann Blandford, Andrew Copas, Paul Flowers, Jo Gibbs, Lynne Haahr, Karen Lloyd, Jennifer MacDonald, Danita Mooney, Nuria Gallego Marquez, Fiona Mapp, Amelia McInnes-Dean, Julie McLeod, Mark Monahan, Jonathan O’Sullivan, Tracy Roberts, Jonathan Ross, John Saunders, Pam Sonnenberg, Merle Symonds, Roos Van Greevenbroek, Andrew Winter, Melvina Woode Owusu*.

## Author contributions

**Funding acquisition**: CSE, PF, JG, MWO, AB, JS, PS; **Conceptualisation**: JMcL, PF, JMacD; **Project administration**: MWO; **Supervision**: PF, CSE; **Methodology**: JMacD, PF; **Resources**: JMcL, JMacD, FM; **Investigation**: JMcL; **Formal analysis**: JMcL, PF; **Visualisation**: JMcL, PF; **Writing – original draft**: CSE, JG, FM, JS, JMcL, PF; **Writing – review & editing**: CSE, JG, FM; JMacD, JMcL, PF, MWO, NGM, AMD, JS, AB, PS.

## Appendix A

Target sampling frame, Socio-economic demographics and screening survey, Interview topic guide, Sexual health support resources, Participant self-reported skills and experience using the internet

## Appendix B

BCW analysis

