## Appendix A for "Recommendations for developing asynchronous online consultations for chlamydia treatment for underserved populations: A Behaviour Change Wheel analysis"

### Target sampling frame

| Demographic characteristics and socioeconomic indicators | | Targets | Final sample [c] |
|---|---|---|---|
| **PROGRESS** | | | |
| **Place of residence (SES [a])** | **Area deprivation (IMD [b])** | n=10-15 from the most deprived quintiles | **n=17** from the most deprived quintiles (n=9 most deprived, n=8 second most deprived) |
| **Race/Ethnicity** | **Ethnicity** | n=10 from minoritised ethnic groups (especially Black Caribbean, Black Other, and Mixed ethnic populations) | **n=9** from minoritised ethnic groups (n=3 Black African, n=1 Mixed/other (Black African and White), n=4 Asian Pakistani, n=1 Asian Chinese) |
| **Occupation (SES [a])** | **Employment** | n=10 out of work / in temporary work | **n=11** out of work |
| **Gender/Sex** | **Gender** | n=10-15 cisgender men | **n=16** cisgender men |
| | | n=10-15 cisgender women | **n=16** cisgender women |
| | | n=5-10 non-binary and trans | n=2 non-binary and trans (n=1 non-binary, n=1 non-binary/transmasc) |
| **Education (SES [a])** | **Education** | n=10-15 with no higher education | **n=18** no higher education (n=6 high school, n=12 college) |
| **Plus** | | | |
| **Age** | **Age** | n=15 aged under 25 years | n=8 aged under 25 years |
| | | n=10 aged 25 to 49 years | **n=23** aged 25-49 |
| | | n=5 aged 50+ years | n= 3 aged 50+ |
| **Sexual orientation/ behaviour/ identity** | **Sexuality** | n=5-10 gay and bisexual men who have sex with men (GBMSM) | **n=9** GBMSM (n=2 bisexual, n=7 gay) |
| | | n=15-20 heterosexual men | n=6 heterosexual men |
| | | n=5 heterosexually identifying men who have sex with men (HSMSM) | n=0 HIMSM |
| | | n=5 bisexual women | **n=6** bisexual women |
| **Disability** | **Disability** | n=5 with a mild learning disability | **n=10** with a learning disability |

### Socio-economic demographics and screening survey

| **Eligibility Questions** |
|---|
| 1. Are you aged 16 or over? |
| 2. Do you live in the UK? |
| 3. Do you have you have access to the internet? |
| 4. Have you had sex in the last 12 months? |
| 5. a) Have you ever ordered a postal STI self-sampling kit before?<br>b) *[If yes]* Did you experience any difficulty in ordering it? |
| **PROGRESS+ screening questions:** |
| 1. What age are you? |
| 2. How would you best describe your gender? |
| 3. Were you assigned a different gender at birth? |
| 4. How would you describe your sexual orientation? |
| 5. How would you describe your ethnicity? |
| 6. What is your highest educational qualification? |
| 7. How would you describe your current occupation? |
| 8. What is your postcode? |
| 9. a) Do you have any physical or mental health conditions or illnesses that have lasted or are expected to last for 12 months or more?<br>b) *[If yes]* Do any of your illnesses or conditions reduce your ability to carry out day to day activities? |
| 10. Do you consider yourself to have a learning disability? |
| **PROGRESS+ demographic questions:** |
| 1. a) Were you born in the UK?<br>b) *[If no]* Which country were you born in? |
| 2. What is your first language? |
| 3. a) Do you regard yourself as belonging to any particular religion or faith?<br>b) *[If yes]* Which religion or faith? |
| 4. Do you ever have difficulty making ends meet at the end of the month? |
| 5. How many people can you call upon to provide support if you need it? |
| **Internet access and use questions:** |
| 1. How do you typically access the Internet? |
| 2. What device(s) do you use to access the internet? |
| 3. Are these your own devices? |
| 4. How often do you go online? |
| 5. What sorts of things do you go online for? |
| 6. a) Have you ever searched for ~~health or~~ sexual health information online?<br>b) *[if yes]* What sort of information have you searched for? |
| 7. a) Have you ever used any online ~~health or~~ sexual health services?<br>b) *[if yes]* Which services have you used?<br>• prompt about live chat/email or text service |
| 8. How would you rate your skills in using the internet? |

### Interview topic guide

| **Behavioural domain**<br>Description of element of online sexual healthcare and corresponding slide | **Specific behaviour** | | **Question** | **Prompts** |
|---|---|---|---|---|
| **Knowledge of current online sexual healthcare (1 min)** | - | | *[If never searched for/used SHS online before]*<br>**Can you briefly tell me about your experience of accessing sexual health services in the past?** | • How did you become aware of these health / sexual health services?<br>• What, if anything, has affected your decision not to use online sexual health services?<br>• What do you think about online sexual health services as an idea?<br>o What do you think it may be like?<br>o What do you think it might involve?<br>• Do you have a good idea of what STIs are? |
| | | | *[If searched for/used SHS online before]*<br>**Can you briefly tell me, from your recent experience, what you know about online sexual health services/ health services?** | • How did you become aware of these online sexual health services?<br>• Can you tell me what has affected your decision to use online sexual health services? |
| **Online sexual healthcare *per se* (2 mins)** | **Access online sexual healthcare** | | **Is there anything that might make/has made it difficult for you to access online sexual health services?** | • Is there anything that might make it difficult for you to get online to use online SHS?<br>• Think about how you normally access the internet |
| | **Use online sexual healthcare** | | **Is there anything that might/has make it difficult for you to use online sexual health services?** | |

| | | | | |
|---|---|---|---|---|
| **1. Getting sexual health information online (5-7 mins)**<br><br>*[#1 on slide 3 & Slide 4]*<br>You can find a wide range of information about sexual health online, on websites like the NHS and sexual health charity websites. These websites have information on them about sexual health services, including clinic locations and opening times. They also have information about sexual health, including how to have safe sex and STI symptoms and testing<br>*[Slide 5]*<br>You can search for this information on Internet, using devices like a mobile phone or computer.<br>*[Sense check]* | - | Previous experience | *[If never searched for sexual health or health information online before]*<br>**Tell me about your experience using the internet for any kind of information** | • What, if anything, has stopped you from searching for health/sexual health information online?<br>• Compared to getting sexual health information in person or on the phone |
| | | | *[If only searched for other health information]*<br>**Tell me about any experience of searching for other health information online** | • What are your thoughts on searching for SH information online? |
| | | | *[If have searched for sexual health information online before]*<br>**Tell me about your previous experience of searching for sexual health information online** | • What are your thoughts on searching for SH information online? |
| | | Barriers | **What might make/ has made it difficult for you to search for sexual health information online?** | • Why?<br>• What might you struggle with?<br>• Is there anything that might stop you/put you off?<br>• Is there anything you might be concerned about?<br>• How would you know if you could trust the website?<br>• What challenges may there be for you in searching for sexual health information online? |

| | | | | |
|---|---|---|---|---|
| | | Facilitators | **What might make/has made it easy for you to search for sexual health information online?** | • Why?<br>• What are the benefits for you to this?<br>• What might encourage you to do this?<br>• What would help you do this?<br>• What do you think would be good about it? Why? |
| **2. Getting sexual health information and advice online (5-7 mins)**<br><br>*[#2 on slide 3 & slide 6]*<br>Some sexual health clinic and charity websites offer a service that lets you talk to a sexual healthcare professional (doctor or nurse on the NHS) or trained staff (on a sexual health charity website). You can ask or talk about things like safe sex, sexually transmitted infections, and the services available to you.<br>These are screenshots of these services from a sexual health website.<br>*[Slide 7]*<br>This includes a live chat service where you type your name and a question and are connected to someone immediately, or an email or text service here you | **-** | | *[If never got sexual health support and advice online before]*<br>**What do you think about getting sexual health support and advice online?** | • What might be different for you about using a live chat and an email or text service to get sexual health support and advice online?<br>• Do you think there might be any difference between talking to a HCP or a trained member of staff |
| | **Live chat** | | *[If never used a live chat for getting sexual health support and advice online before]*<br>**What do you think about using a live chat service to get sexual health support and advice online?** | • Why? |
| | | Barriers | **What would make/ has made it difficult for you to use a live web chat to get sexual health support and advice online?** | • Is there anything you might be concerned about?<br>• Is there anything that would stop you?<br>• What might put you off using a live web chat for getting sexual health support and advice?<br>• Why wouldn't you do this?<br>• What challenges might there be for you?<br>• What might help you with this? |

| | | | | |
|---|---|---|---|---|
| type your name, email address or phone number and a question, and they respond to you by email or text as soon as possible (generally within a few hours to days)<br>*[Sense check]* | | Facilitators | **What would make/ has made it easy for you to use a live web chat to get sexual health support and advice online?** | • What do you think would encourage you to use a live web chat for getting sexual health support and advice?<br>• Why would you do this?<br>• Difference between talking to HCP and trained staff? |
| | **Email/Text service** | | *[If never used an email/text service for getting sexual health support and advice online before]*<br>**What do you think about using an email/text service to get sexual health support and advice online?** | • Why? |
| | | Barriers | **What would make/ has made it difficult for you to use an email/text service to get sexual health support and advice online?** | • What might stop you?<br>• What might put you off using an email/text service for getting sexual health support and advice?<br>• Why wouldn't you do this?<br>• What challenges might there be?<br>• What might help you with this? |
| | | Facilitators | **What would make/ has made it easy for you to use an email or text service to get sexual health support and advice online?** | • What would encourage you to use an email/text service to get sexual health support and advice?<br>• Why would you do this? |
| | **Sexual health service** | Barriers | | • Is there anything that would make it difficult for you to collect your STI treatment from a pharmacy?<br>• Challenges for you |
| | | Facilitators | | • Is there anything that would make it easy for you to collect your STI treatment from a SHS?<br>• Benefits for you |

### Sexual health support resources

**Sources of support**

We want to make sure that you have a list of services that offer different types of support and information in case something came up in the interview that was difficult or uncomfortable for you.

**Sexual health services and organisations**

| Name and details of support offered | Website and contact details |
|---|---|
| *Sandyford Sexual Health Service* offers sexual healthcare for those living in the Glasgow area | https://sandyford.scot \| 0141 211 8130 \| |
| *The Chalmers Centre* offers sexual healthcare for those living in the Lothian area | https://lothiansexualhealth.scot \| 0131 536 1070 |
| To find your nearest sexual health service visit: | https://sexualhealthscotland.co.uk/get-help/sexual-health-service-finder |
| *S-X* provide information on sexual health, relationships, and mental health for gay and bisexual men, and all men who have sex with men living in Scotland (including testing services) | https://s-x.scot \| 0131 652 3250 or 07703 840970 \| |
| *Terrence Higgins Trust* provide information on sexual health, safer sex, and sexually transmitted infections, including testing services | https://tht.org.uk/sexual-health \| 0808 802 1221 (direct helpline) |
| *Waverley Care* provide information on sexual health, sex, and relationships | https://www.waverleycare.org/support-and-advice/sexual-health |
| *The African Health Project* provide support and advice on HIV and sexual health (and a range of other health and social care issues) | 0141 332 2520 \| 0131 558 1425 |
| *Young Scot* offer information about sex, sexual health, and relationships for young people living in Scotland | https://young.scot/campaigns/national/sexual-health |

**Wider health services and organisations**

| Name and details of support offered | Website and contact details |
|---|---|
| *NHS Inform* provide up-to-date information about COVID-19 | https://www.nhsinform.scot/illnesses-and-conditions/infections-and-poisoning/coronavirus-covid-19 |
| *NHS 24* provide urgent care advice and mental health support | https://www.nhs24.scot \| 111 |
| *Breathing Space* have experienced advisors who will listen and offer information and advice regarding mental health | https://breathingspace.scot \| 0800 83 85 87 |
| *Hwupenyu Health and Wellbeing Project* offer information, support, and guidance on health, mental health, and social topics for Black ethnic communities living in Scotland | http://www.hwupenyuproject.org/ \| 0141 418 0940 \| |
| *The Scottish Refugee Council* provide practical support, advice, and a listening ear for people in need of refugee protection | https://www.scottishrefugeecouncil.org.uk/contact-us/ \| 0808 196 7274 |
| *Switchboard* provide an information, support and referral service for LGBTQ+ people | https://switchboard.lgbt/ \| 0300 330 0630 \| |
| *Disability Information Scotland* provide reliable, accurate, and accessible information for people living with disability in Scotland | https://www.disabilityscot.org.uk/ \| 0300 323 9961 |
| *Victim Support Scotland* provide support and information to people affected by crime, including but not limited to domestic abuse and sexual assault | https://victimsupport.scot \| 0800 160 1985 |

### Participant self-reported skills and experience using the internet

| Variables | n | % [a] |
|---|---|---|
| **Digital literacy** | | |
| **Skills using the internet** | | |
| High (8-10; pretty good, confident, excellent) | 20 | 57.1 |
| Medium (4-7; not great, getting better, not bad) | 12 | 34.3 |
| Low (0-3) | 2 | 5.7 |
| **Experience using the internet for sexual health** | | |
| **Previously searched for health or sexual health information** | | |
| Yes | 20 | 57.1 |
| No | 11 | |
| Yes, health only, not sexual health | 2 | 5.7 |
| Yes, tried and struggled | 1 | 2.9 |
| **Sexual health information searched for** | | |
| Sexually transmitted infections (STIs)/ blood borne viruses (BBV) symptoms (including yeast infection, thrush, urinary tract infections) | 11 | 31.4 |
| Information about STIs/BBV (including most common, how they're contracted) | 7 | 20.0 |
| How/where to get tested | 5 | 14.3 |
| Where to find local clinics | 3 | 8.6 |
| Information about contraception (including the coil, morning after pill) | 3 | 8.6 |
| Information about HIV (including support groups, pre-exposure prophylaxis) | 2 | 5.7 |
| Treatment for STIs/HIV (e.g., what the options are, where to get treatment) | 2 | 5.7 |
| Window periods | 2 | 5.7 |
| Efficacy and reliability of tests | 1 | 2.9 |
| Trans specific sexual health | 1 | 2.9 |
| Sexual health in general | 1 | 2.9 |
| **Previous use of online sexual health services** | | |
| Live chat or email/text exchange service | 0 | 0 |
| Booking appointment for in-person clinic online | 4 | 11.4 |
| Ordering medication (private clinic) | 1 | 2.9 |
| **Ordered or used an online postal STI/BBV self-sampling (OPSS) kit** | | |
| Never ordered an OPSS | 24 | 68.6 |
| Ordered and struggled to use an OPSS (e.g., blood sampling) | 7 | 20.0 |
| Struggled to order an OPSS | 3 | 8.6 |
| **Experience using the internet** | | |
| **Devices used to access the internet** | | |
| Laptop/computer and/or mobile phone | 20 | 57.1 |
| Phone only | 6 | 17.1 |
| Multiple devices (e.g., smart TV, mobile phone, tablet, computer, laptop) | 4 | 11.4 |
| Phone and tablet | 2 | 5.7 |
| Tablet only | 1 | 2.9 |
| iPad | 1 | 2.9 |
| WiFi | 1 | 2.9 |

| | | |
|---|---|---|
| **Own device owned** | | |
| Yes | 30 | 85.7 |
| Yes, phone only | 3 | 8.6 |
| No | 1 | 2.9 |
| **How often online** | | |
| Every day/daily | 25 | 71.4 |
| Constantly/all the time/every hour | 4 | 11.4 |
| Weekly or monthly | 2 | 5.7 |
| Not often/not too much | 2 | 5.7 |
| Once or twice a week | 1 | 2.9 |
| **What participants go online for** | | |
| Social media (e.g., Facebook, Instagram, Twitter, WhatsApp) | 22 | 62.9 |
| School/University work (e.g., homework, research, classwork/coursework) | 12 | 34.3 |
| TV streaming (e.g., YouTube, Netflix) | 10 | 28.6 |
| Engine searches (e.g., how to spell words, "NHS searches", google images) | 9 | 25.7 |
| Online shopping | 7 | 20.0 |
| News | 6 | 17.1 |
| Work (e.g., rotas, research) | 5 | 14.3 |
| Checking emails | 5 | 14.3 |
| Online groups (e.g., prayer group, Zoom meetings) | 3 | 8.6 |
| Contacting friends/family | 3 | 8.6 |
| Online gaming | 2 | 5.7 |
| Steaming music | 2 | 5.7 |
| Entertainment | 1 | 2.9 |
| Booking appointments | 1 | 2.9 |
| Online banking | 1 | 2.9 |
| Meditation/motivational speeches | 1 | 2.9 |
| "Random" | 1 | 2.9 |

*Participant demographics for one participant were not obtained, table includes demographics for n=34, except where participants did not wish to answer the question. Percentages are calculated for N=35; those not summing to 100 are due to rounding. ** Responses to the question "how would you rate your skills using the internet?" were highly varied. Here, all response were categorised into 'high' ("eight" to "ten", "great", "pretty/very/quite good", "quite high", "excellent", "confident"), 'medium' ("four" to "seven", "not great", "not bad", "not too sure, I'm still learning", "getting better", "medium", "couldn't do anything fancy"), and 'low' ("zero" to "three", "low").
