## Appendix B for "Recommendations for developing asynchronous online consultations for chlamydia treatment for underserved populations: A Behaviour Change Wheel analysis"

### Intervention content and recommendations from Behaviour Change Wheel to address barriers and facilitators to accessing and using Asynchronous online consultations

| **Barrier/ Facilitator theme** | **Barrier/ Facilitator sub-theme** | **Barrier**<br>**People from populations that might struggle using online SHS** would find it **difficult** to complete an automated online consultation to get STI treatment… | **Facilitator**<br>**People from populations that might struggle using online SHS** would find it **easy** to complete an automated online consultation to get STI treatment... | **COM-B** | **TDF** | **Intervention functions** | **BCT** | **Recommendation**<br>To improve [behavioural domain] for [target], [actor] should **[action], at/on [point in the pathway - time/context]** to address [barrier/facilitator]. |
|---|---|---|---|---|---|---|---|---|
| 1 Awareness and familiarity | Familiarity with online healthcare | due to lack of experience with online SHC and familiarity with in-person treatment | N/A | Reflective motivation | Beliefs about capabilities | Modelling | Demonstration of the behaviour (6.1) | Demonstrate to patients how the online consultation works, what is involved and how to complete it |
| 1 Awareness and familiarity | Familiarity with online healthcare | due to lack of experience with online SHC and familiarity with in-person treatment | N/A | Reflective motivation | Beliefs about capabilities | Enablement | Graded tasks (8.7) | Allow and ask the patient to complete the form in stages - starting with the easiest information (e.g., personal info) to the most difficult (e.g., health info) |
| 1 Awareness and familiarity | Familiarity with online healthcare | due to lack of experience with online SHC and familiarity with in-person treatment | N/A | Reflective motivation | Beliefs about capabilities | Persuasion | Verbal persuasion about capability (15.1) | Tell the person that they can successfully complete the online consultation, despite the lack of familiarity |

| | | | | | | | | |
|---|---|---|---|---|---|---|---|---|
| 1 Awareness and familiarity | Awareness of the service | N/A | if they know of the service | Psychological capability | Knowledge | Education | Prompts/cues (7.1) | Advertise the online consultation on the eSHC - provide a link to the online consultation from the results page |
| 1 Awareness and familiarity | Awareness of the service | N/A | if they know of the service | Physical opportunity | Environmental context and resources | Environmental restructuring | Adding objects to the environment (12.5) | Advertise the online consultation in offline spaces, such as in-person clinics, GP - in leaflet or poster form |
| 2 Perceived needs | Perceive the need to see a healthcare professional in person | due to perceived need to see HCP in person for reassurance, to ask questions or get examination | N/A | Reflective motivation | Beliefs about consequences | Persuasion | Framing/ reframing (13.2) | Suggest to patients that they think of the online consultation as increasing sexual healthcare self-management and independence (rather than reducing their ability to see a HCP) |
| 2 Perceived needs | Perceive the need to see a healthcare professional in person | due to perceived need to see HCP in person for reassurance, to ask questions or get examination | N/A | Reflective motivation | Beliefs about consequences | Persuasion | Verbal persuasion about capability (15.1) | Tell patients that they can successfully complete the online consultation and get treatment without the need for an examination or providing extra details - i.e., form has been created so this is not necessary |
| 2 Perceived needs | Perceived level of risk of an STI is low | if level of perceived risk is high (e.g., HIV, have symptoms) | if level of perceived risk is low (e.g., no symptoms, bacterial STI) | Reflective motivation | Beliefs about consequences | Persuasion | Framing/ reframing (13.2) | Suggest to patients that they think of the online consultation as increasing sexual healthcare self-management and independence for when they have symptoms (rather than reducing their ability to have symptoms checked by HCP) |

| | | | | | | | | |
|---|---|---|---|---|---|---|---|---|
| 2 Perceived needs | Perceive the need to see a healthcare professional in person | due to perceived need to see HCP in person for reassurance, to ask questions or get examination | N/A | Reflective motivation | Beliefs about consequences | Persuasion | Information about health consequences (5.1) | Tell patients that not being examined/not seeing an HCP in person is **not** detrimental to their health - that they will receive the same treatment through completing online form as they would if they saw a HCP in person |
| 2 Perceived needs | Perceived level of risk of an STI is low | if level of perceived risk is high (e.g., HIV, have symptoms) | if level of perceived risk is low (e.g., no symptoms, bacterial STI) | Reflective motivation | Beliefs about consequences | Education | information about health consequences (5.1) | Inform patients online consultation is for treatment for a bacterial STI and that online consultation is designed to help people get treatment for symptoms to reduce consequences of having an STI |
| 2 Perceived needs | Perceived level of risk of an STI is low | if level of perceived risk is high (e.g., HIV, have symptoms) | if level of perceived risk is low (e.g., no symptoms, bacterial STI) | Reflective motivation | Beliefs about consequences | Persuasion | Credible source (9.1) | Provide a video of a speech given by a trusted HCP (Claudia?) explaining the robust research that went into developing the eSHC to ensure its safety and ability to appropriately treat someone with symptoms, as well as its benefits (e.g., anonymity, convenience etc.) |
| 3 Convenience and Resources | Increase convenience and control | N/A | because it is (perceived to be) more convenient than in-person services and gives control over time | Reflective motivation | Beliefs about consequences | Education | Information about social and environmental consequences (5.3) | Tell patients that the online consultation can increase their control over the time they can request and access treatment |
| 3 Convenience and Resources | Increase convenience and control | N/A | because it is perceived to be faster than in person services | Reflective motivation | Beliefs about consequences | Persuasion | Information about social and environmental consequences (5.3) | Tell patients that the online consultation can increase speed of getting treatment |

| | | | | | | | | |
|---|---|---|---|---|---|---|---|---|
| 4 Privacy | Concerned about others reading responses | due to concerns about others finding out | because it is private and anonymous | Reflective motivation | Beliefs about consequences | Persuasion | Information about social and environmental consequences (5.3) | Inform patients that it is entirely anonymous and confidential - personal information does not get released to anyone, including GP, family etc. |
| 4 Privacy | Perceived to be private | N/A | because it is perceived to offer protection from negative experiences in clinic/with HCP | Reflective motivation | Beliefs about consequences | Education | Information about social and environmental consequences (5.3) | Inform patients that the online consultation is entirely self-managed and allows them to take getting treatment into their own hands without having to see a HCP |
| 4 Privacy | Concerned about others reading responses | due to concerns about others finding out | because it is private and anonymous | Social opportunity | Social influences | Enablement | Pros and cons (9.2) | Get patients to think about the pros and cons of the online consultation for them, e.g., completely anonymous, more convenient than in person appointment) vs concern about others seeing them complete the form) |
| 5 Answering personal questions online | Questions are perceived to be necessary and expected | due to discomfort answering personal questions (concerns about discrimination, judgement, anonymity, faith, gender, & perception of necessity) | because questions are perceived to be expected and necessary to get correct treatment | Automatic motivation | Emotion | Enablement | Framing/ reframing (13.2) | Suggest to patients that they think of the personal questions in the online consultation as necessary to get correct treatment (rather than uncomfortable and too personal) |
| 5 Answering questions correctly | Understanding or interpreting the questions | due to difficulty reading and understanding (learning difficulty) | if it is easy-read with pictures | Physical opportunity | Environmental context and resources | Environmental restructuring | Restructuring the physical [digital environment (12.1) | Ensure the form is easy-read: short, simple sentences; no medical jargon (or medical jargon explain very simply); provide pictures to enable understanding of questions and response options. Provide audio option for reading out questions and response options. |

| | | | | | | | | |
|---|---|---|---|---|---|---|---|---|
| 5 Answering questions correctly | Understanding or interpreting the questions | due to concerns about getting incorrect treatment due to a) misunderstanding /misinterpreting questions; b) lack of trust in computer system; c) inability/ not qualified to identify health issues; d) concern about completing form incorrectly (putting wrong information in | N/A | Reflective motivation | Beliefs about consequences | Education | Prompts/cues (7.1) | Provide an 'unsure about this question/which response applies to you? Click here for more details/information'. If they click this, provide extra information to help interpret the question and written encouragement about their ability to interpret questions correctly. |
| 5 Answering questions correctly | Having support to complete it | N/A | if they have options for support from HCP | Physical opportunity | Environmental context and resources | Environmental restructuring | Adding objects to the environment (12.5) | Provide options for support for completing the online consultation throughout - phone number(s) to call; live chat |
| 5 Answering questions correctly | Having support to complete it | N/A | if they have options for support from HCP | Physical opportunity | Environmental context and resources | Environmental restructuring | Restructuring the physical environment (12.1) | Ensure the options to contact for help (e.g., clinical hotline, live chat etc.) are highlighted, clear and easy to see throughout the online consultation |
| 5 Answering questions correctly | Having support to complete it | N/A | if they have options for support from HCP | Physical opportunity | Environmental context and resources | Enablement | Social support (practical) (3.2) | Provide options for support for completing the online consultation - phone number(s) to call; live chat |
| 5 Answering questions correctly | Layout and questions of the consultation simple and clear | N/A | if the layout and questions are straightforward, simple and clear (step-by-step) | Physical opportunity | Environmental context and resources | Training | Instruction on how to perform a behaviour (4.1) | Provide step-by-step instruction on how to complete the online consultation |

| | | | | | | | | |
|---|---|---|---|---|---|---|---|---|
| 5 Answering questions correctly | Understanding or interpreting the questions | due to concerns about getting incorrect treatment due to a) misunderstanding /misinterpreting questions; b) lack of trust in computer system; c) inability/ not qualified to identify health issues; d) concern about completing form incorrectly (putting wrong information in | N/A | Reflective motivation | Beliefs about capabilities | Modelling | Demonstration of the behaviour (6.1) | Demonstrate to patients how to complete the form correctly e.g., show examples of completed forms |
| 5 Answering questions correctly | Understanding or interpreting the questions | due to concerns about getting incorrect treatment due to a) misunderstanding /misinterpreting questions; b) lack of trust in computer system; c) inability/ not qualified to identify health issues; d) concern about completing form incorrectly (putting wrong information in | N/A | Psychological capability | Cognitive skills | Training | Instruction on how to perform a behaviour (4.1) | Provide initial brief explanation of the task - what the automated online consultation involves and how to complete the form. |
| 5 Answering questions correctly | Understanding or interpreting the questions | due to difficulty reading and understanding (learning difficulty) | if it is easy-read with pictures | Psychological capability | Cognitive skills | Training | Instruction on how to perform a behaviour (4.1) | Provide demonstrations of this, e.g., short videos or pictures of examples |
| 5 Answering questions correctly | Understanding or interpreting the questions | due to difficulty reading and understanding (learning difficulty) | if it is easy-read with pictures | Psychological capability | Cognitive skills | Training | Demonstration of the behaviour (6.1) | Provide initial brief explanation of the task - what the automated online consultation involves and how to complete the form. Provide demonstrations of this, e.g., short videos or pictures of examples |

| | | | | | | | | |
|---|---|---|---|---|---|---|---|---|
| 5 Answering questions correctly | Layout and questions of the consultation simple and clear | N/A | if the layout and questions are straightforward, simple and clear (step-by-step) | Physical opportunity | Environmental context and resources | Training | Graded tasks (8.7) | Allow and ask the patient to complete the form in stages - starting with the easiest information (e.g., personal info) to the most difficult (e.g., health info) |
| 5 Answering questions correctly | Understanding or interpreting the questions | due to difficulty reading and understanding (learning difficulty) | if it is easy read with pictures | Psychological capability | Cognitive skills | Training | Graded tasks (8.7) | Allow and ask the patient to complete the form in stages (over time?) - starting with the easiest information (e.g., personal info) to the most difficult (e.g., health info) |
| 5 Answering questions correctly | Layout and questions of the consultation simple and clear | N/A | if the layout and questions are straightforward, simple and clear (step-by-step) | Physical opportunity | Environmental context and resources | Environmental restructuring | Restructuring the physical environment (12.1) | Ensure the layout of the online consultation is simple, straightforward, and step by step - i.e., in small 'chunks' to be completed; clear simple headings; page(s) not busy, only questions and responses |
| 5 Answering questions correctly | Understanding or interpreting the questions | due to concerns about getting incorrect treatment due to a) misunderstanding /misinterpreting questions; b) lack of trust in computer system; c) inability/ not qualified to identify health issues; d) concern about completing form incorrectly (putting wrong information in | N/A | Reflective motivation | Beliefs about capabilities | Persuasion | Verbal persuasion about capability (15.1) | Tell patients that they can successfully complete the form correctly - provide a couple of short sentences to encourage people regarding their ability to correctly interpret the questions and identify health concerns (e.g., deep pain after sex and skin lumps). Throughout the form, provide written encouragement about their ability to interpret questions correctly and self-identify health issues |
| 5 Answering questions correctly | Understanding or interpreting the questions | due to concerns about getting incorrect treatment due to a) misunderstanding /misinterpreting questions; b) lack of trust in computer system; c) inability/ not qualified to identify health issues; d) concern about completing form | N/A | Reflective motivation | Beliefs about consequences | Education | Information about social and environmental consequences (5.3) | Explain that the patient will not get incorrect treatment - if they complete the form 'incorrectly', they will be referred to in person services |

| | | | | | | | | |
|---|---|---|---|---|---|---|---|---|
| | | incorrectly (putting wrong information in | | | | | | |
| 5 Answering questions correctly | Understanding or interpreting the questions | due to concerns about getting incorrect treatment due to a) misunderstanding /misinterpreting questions; b) lack of trust in computer system; c) inability/ not qualified to identify health issues; d) concern about completing form incorrectly (putting wrong information in | N/A | Reflective motivation | Beliefs about capabilities | Persuasion | Credible source (9.1) | Present a speech given by trusted HCP on how the online consultation was developed and how it works - robust research to create it - to address lack of trust in the system |
| 6 Answering personal questions online | Uncomfortable answering personal questions | due to discomfort answering personal questions (concerns about discrimination, judgement, anonymity, faith, gender, & perception of necessity) | because questions are perceived to be expected and necessary to get correct treatment | Reflective motivation | Beliefs about consequences | Persuasion | Information about social and environment consequences (5.3) | Inform Muslim participants that online consultation is fully automated, so they will not be sharing sexual information with another person |
| 6 Answering personal questions online | Uncomfortable answering personal questions | due to discomfort answering personal questions (concerns about discrimination, judgement, anonymity, faith, gender, & perception of necessity) | because questions are perceived to be expected and necessary to get correct treatment | Automatic motivation | Emotion | Enablement | Reduce negative emotions (11.2) | Reduce discomfort about completing personal questions by providing reassurance about anonymity and no of judgement or discrimination |
| 6 Answering personal questions online | Questions are perceived to be necessary and expected | due to discomfort answering personal questions (concerns about discrimination, judgement, anonymity, faith, gender, & perception of necessity) | because questions are perceived to be expected and necessary to get correct treatment | Automatic motivation | Emotion | Enablement | Pros and cons (9.2) | Highlight the pros of completing the online consultation (getting treatment) over the cons (answering personal questions) and ask patients to weigh up the pros and cons for themselves |

| | | | | | | | | |
|---|---|---|---|---|---|---|---|---|
| 6 Answering personal questions online | Uncomfortable answering personal questions | due to discomfort answering personal questions (concerns about discrimination, judgement, anonymity, faith, gender, & perception of necessity) | because questions are perceived to be expected and necessary to get correct treatment | Reflective motivation | Beliefs about consequences | Education | information about health consequences (5.1) | Inform patients that if online form is not completed, they will not get treatment, emphasising consequences of leaving an STI untreated |
| 6 Answering personal questions online | Questions are perceived to be necessary and expected | due to discomfort answering personal questions (concerns about discrimination, judgement, anonymity, faith, gender, & perception of necessity) | because questions are perceived to be expected and necessary to get correct treatment | Automatic motivation | Emotion | Enablement | Comparative imagining of future outcomes (9.3) | Prompt the person to imagine and compare likely or possible outcomes following not completing the personal questions versus completing the form |
| 6 Answering personal questions online | Uncomfortable answering personal questions | due to discomfort answering personal questions (concerns about discrimination, judgement, anonymity, faith, gender, & perception of necessity) | because questions are perceived to be expected and necessary to get correct treatment | Reflective motivation | Beliefs about consequences | Education | Information about social and environment consequences (5.3) | Inform patients that questions asked are the same questions they would be asked in person, highting that questions will be required to be answered to get treatment |
